# Druggable causal genes of bipolar disorder identified through Mendelian Randomization analysis offer a route to intervention in integrated stress response

**DOI:** 10.1101/2023.12.20.23300345

**Authors:** Zhai Ting

## Abstract

Bipolar disorder (BD) is known to be comorbid with many medical and psychiatric conditions and they may share inflammatory and stress-related aetiologies, which could give rise to this association. Psychosomatic illnesses are characterized by widespread immune signaling in the central nervous system, oxidative stress, and increased immune trafficking into the brain. The integrated stress response (ISR) responds to a variety of different stress conditions that lead to alterations in cellular homeostasis. However, as a causative mechanism underlying the cognitive deficits and neurodegeneration in a broad range of brain disorders, the impact of ISR on BD is understudied, particularly for the ISR’s central regulatory switch lies in eIF2 ternary complex formation. We aimed to prioritize eIF2-associated genes for follow-up studies using the summary data-based Mendelian Randomization (SMR) and Bayesian colocalization (COLOC) methods to integrate the summary-level data of the GWAS on BD and the expression quantitative trait locus (eQTL) study in blood. We utilized the GWAS data including 41917 BD cases and 371549 controls from the Psychiatric Genomics Consortium and the eQTL data from 31,684 participants of predominantly European ancestry from the eQTLGen consortium. Plus, we use Graph DB to alt drug targets. The SMR analysis identified EIF2B5 gene that were associated with BD due to no linkage but pleiotropy or causality. The COLOC analysis strongly suggested that EIF2B5 and the trait of BD were affected by shared causal variants, and thus were colocalized. Utilizing data in EpiGraphDB we find other putative causal BD genes (EIF2AK4 and GSK3B) to prioritise potential alternative drug targets.

## 1 Introduction

Bipolar disorders(In this article, the abbreviation BD will be used to refer to Bipolar disorders) are described by the *Diagnostic and statistical manual of mental disorders: DSM-5™, 5th ed* (2013) as a group of brain disorders that cause extreme fluctuation in a person’s mood, energy, and ability to function. People who live with bipolar disorder experience periods of great excitement, overactivity, delusions, and euphoria (known as mania) and other periods of feeling sad and hopeless (known as depression). The prevalence of bipolar disorders was consistent across diverse cultures and ethnic groups, with an aggregate lifetime prevalence of 0.6% for bipolar I disorder, 0.4% for bipolar II disorder, 1.4% for subthreshold bipolar disorder, and 2.4% for the bipolar disorder spectrum(Grande et al., 2016). A better understanding of the underlying pathophysiology may aid in improving diagnostic accuracy and patient treatment stratification(Vieta et al., 2018); however, the pathophysiology mechanism of BD remains unclear.

As noted by Rowland and Marwaha (2018), while numerous genetic and environmental risk factors have been identified, the attributable risk of individual factors is often small, and most are not specific to bipolar disorder but are associated with several mental illnesses. This is evident in the case of several studies, which have postulated a convergence between BD and medical comorbidities and may point towards shared inflammatory pathophysiology of the disorders(Leboyer et al., 2012; Rosenblat & McIntyre, 2015).

Thus far, a number of studies have revealed a correlation between BD and diverse stress inputs— including proteostasis defects(Mayer et al., 2018), nutrient deprivation(Zhuang et al., 2009), viral infection(Prusty et al., 2018), and redox imbalance(Gu et al., 2015; Kulak et al., 2013; Lackovic et al., 2013). All of these will lead to the reduction in general translation initiation rates and the increase in translation of specific messenger RNAs. These two processes reprogram gene expression to maintain cellular equilibrium, sustain protein-folding capacity, support differentiation, and respond to injury. When the stress cannot be mitigated, the integrated stress response (ISR) triggers apoptosis to eliminate the damaged cell.

The ISR is an evolutionarily conserved intracellular signaling network that helps the cell, tissue, and organism to adapt to a variable environment and maintain health. In response to different environmental and pathological conditions, including protein homeostasis (proteostasis) defects, nutrient deprivation, viral infection, and oxidative stress, the ISR restores balance by reprogramming gene expression. The various stresses are sensed by four specialized kinases (PERK, GCN2, PKR and HRI) that converge on phosphorylation of a single serine on the eukaryotic translation initiation factor eIF2. eIF2 phosphorylation blocks the action of eIF2’s guanine nucleotide exchange factor termed eIF2B, resulting in a general reduction in protein synthesis. However, if the stress cannot be mitigated, the ISR triggers apoptosis to eliminate the damaged cell(Costa-Mattioli & Walter, 2020). Extensive research has shown that the ISR is activated in a wide range of disorders of the brain. This activation is evidenced by detection of eIF2-P and phosphorylation of PKR, PERK, and GCN2 in post-mortem brains from individuals and animal models of cognitive and neurodegenerative disorders, including Alzheimer’s disease, Parkinson’s disease, Huntington disease, amyotrophic lateral sclerosis (ALS), traumatic brain injury, Down syndrome, and Charcot-Marie-Tooth disease (Atkin et al., 2008; Buffington et al., 2014; Chang et al., 2002; Hoozemans et al., 2007; Ishimura et al., 2016; Moon et al., 2018; Smith & Mallucci, 2016). Notably, ISR activation causes cognitive defects in mouse models of traumatic brain injury (Chou et al., 2017; Sen et al., 2017), and Alzheimer’s disease (Hwang et al., 2017; Tible et al., 2019). Whilst increasing data highlight that highly debilitating stress comorbidities such as depression, anxiety disorders, PTSD and epilepsy share pathogenic mechanisms with stress dysfunction and between each other. These mechanisms are probably deeply connected and the structural and functional change caused by one disease triggers the other, despite these it is still not clear on the relationship between them(de Kloet, Joëls, & Holsboer, 2005; de Kloet, Sibug, et al., 2005; Gold et al., 1988; Kanner, 2012).

However, in response to diverse stress stimuli, eukaryotic cells activate a common adaptive pathway, termed the ISR, to restore cellular homeostasis. So, there’s an obvious question, research to date has not yet determined if ISR would bring patients mood disorder, the evidence for this relationship is inconclusive. Considering the negative impacts that multimorbidity of BD can have on people’s life, identifying this multimorbidity patterns can help provide clues to disease prevention and treatment and improve prognosis.

The approach to empirical research adopted for this study was Mendelian Randomization(MR) analysis, which is a reliable method to determine causal effects between risk factors and outcomes using genetic data, which can effectively eliminate the interference of confounding factors (Emdin et al., 2017). In addition, colocalization is an essential analytical method for exploring the common causal molecular mechanism among different diseases and disease-related intermediate phenotypes (Wu et al., 2019). The specific objective of this study was to describe the association between ISR and BD. As a result, we prioritized several eIF2-associated genes for follow-up functional studies in future to elucidate the underlying disease mechanisms. we systematically applied summary data-based Mendelian randomization analysis (SMR) and colocalization (COLOC) to prioritize putative causal genes using publicly available eQTL datasets. Furthermore, we try to explore potential alternative drug targets by searching candidate genes available from EpiGraphDB. Then, using MR analysis to enhance our confidence in identifying potential viable drug targets.

## 2 Methods

### 2.1 Study design and data resources

Figure 1 describes the design of this study.

**Figure 1.**
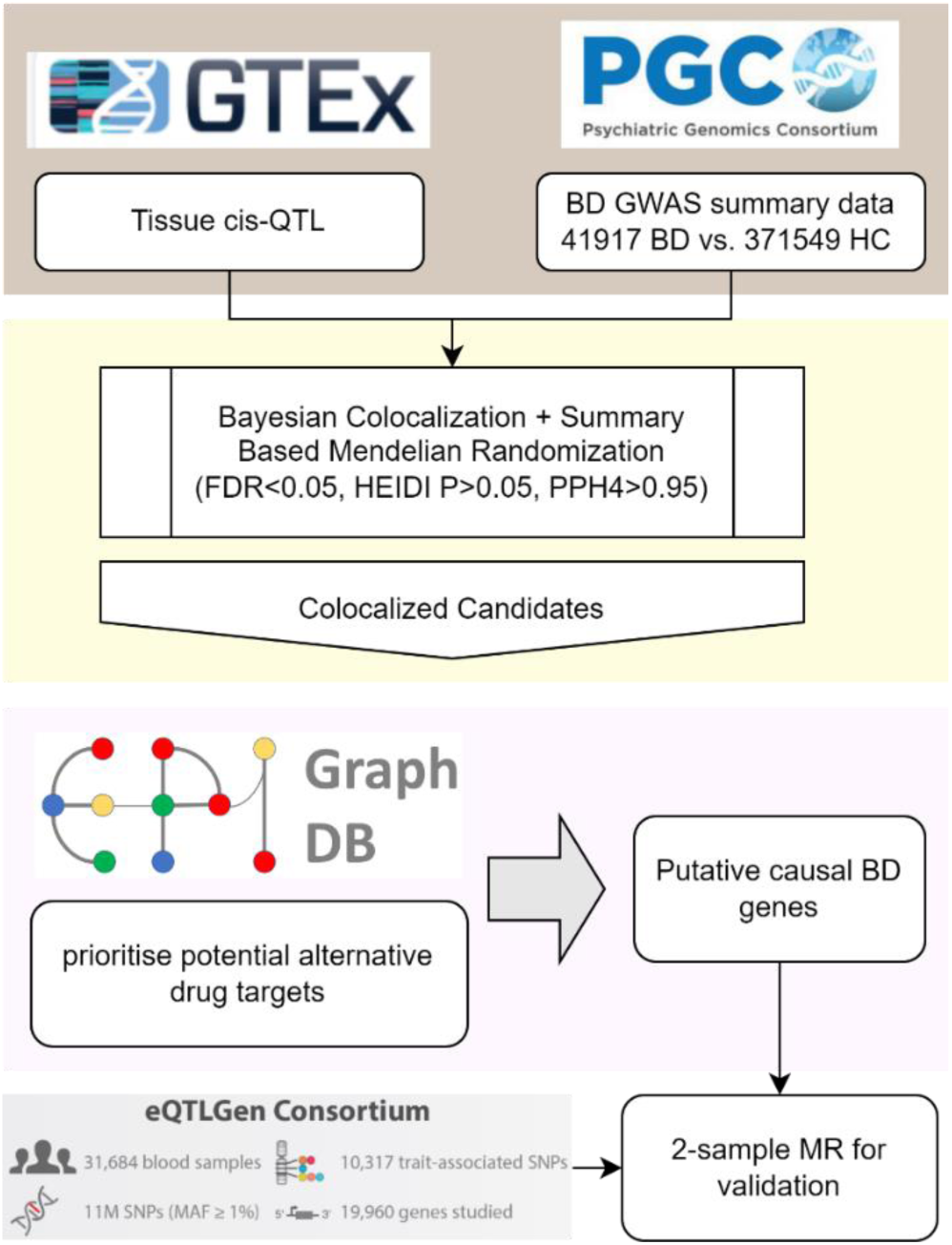
Study overview.

For the summary-level data of the GWAS on BD, we used summary statistics of 41917 BD cases and 371549 controls from the Psychiatric Genomics Consortium (Sullivan, 2023). Tissue cis-QTL data were from the Genotype-Tissue Expression (GTEx) project (Consortium et al., 2020). We downloaded lite version of the GTEx V8 eqtl data (only SNPs with P < 1e-5 are included). This is a set of cis-eQTL summary data across 49 human tissues from the GTEx project. Only SNPs within 1Mb of the transcription start site are available. Blood eQTL summary statistics of eIF2 genes were obtained from eQTLGen including the genetic data of blood gene expression in 31,684 individuals derived from 37 datasets (Võsa et al., 2021).

### 2.2 SMR analysis

We conducted SMR and heterogeneity in dependent instruments (HEIDI) tests in cis regions using the SMR software tool version 1.3.1. Detailed methods for SMR analysis were described in the original SMR paper (Zhu et al., 2016). The SMR software tool was originally developed to implement the SMR & HEIDI methods to test for pleiotropic association between the expression level of a gene and a complex trait of interest using summary-level data from GWAS and expression quantitative trait loci (eQTL) studies. The SMR & HEIDI methodology can be interpreted as an analysis to test if the effect size of a SNP on the phenotype is mediated by gene expression. This tool can therefore be used to prioritize genes underlying GWAS hits for follow-up functional studies.

The default p-value threshold to select the top associated eQTL for the SMR test is 5.0e-8. By default, we only run the SMR analysis in the cis regions. The default threshold of eQTL p-value to select eQTLs for the HEIDI test is 1.57e-3, which is equivalent to a chi-squared value (df=1) of 10. The window centred around the probe to select cis-eQTLs (passing a p-value threshold) for the SMR analysis is 2000Kb.

### 2.3 COLOC analysis

We conducted COLOC analysis using the coloc package (Giambartolomei et al., 2014) in R software (version 4.3.2). The coloc package can be used to perform genetic colocalisation analysis of two potentially related phenotypes, to ask whether they share common genetic causal variant(s) in a given region. This package(version 5) supercedes previously published version 4 by introducing use of the SuSiE approach to deal with multiple causal variants rather than conditioning or masking(Wang et al., 2020).

COLOC analysis calculates posterior probabilities (PPs) of the five hypotheses: 1) H0; no association with either gene expression or phenotype; 2) H1; association with gene expression, not with the phenotype; 3) H2; association with the phenotype, not with gene expression; 4) H3; association with gene expression and phenotype by independent SNVs; and 5) H4; association with gene expression and phenotype by shared causal SNVs. A large PP for H4 (PP.H4 above 0.95) strongly supports shared causal variants affecting both gene expression and phenotype.

### 2.4 PPI networks & Mendelian randomization

Systematic MR of molecular phenotypes such as proteins and expression of transcript levels offer enormous potential to prioritise drug targets for further investigation. However, many genes and gene products are not easily druggable, so some potentially important causal genes may not offer an obvious route to intervention.

A parallel problem is that current GWASes of molecular phenotypes have limited sample sizes and limited protein coverages. A potential way to address both these problems is to use protein-protein interaction information to identify druggable targets which are linked to a non-druggable, but robustly causal target. Their relationship to the causal target increases our confidence in their potential causal role even if the initial evidence of effect is below our multiple-testing threshold.

Here, we use data in EpiGraphDB to prioritise potential alternative drug targets in the same PPI network, as follows: For an existing drug target of interests, we use PPI networks to search for its directly interacting genes that are evidenced to be druggable.(Liu et al., 2020). We then examine the causal evidence of these candidate genes on the disease by using two-sample Mendelian randomization (MR) (Hemani et al., 2018).

## 3 Results

### 3.1 SMR and COLOC analyses using blood eQTL datas

The overall results are presented in Table 1.

**Table 1.**
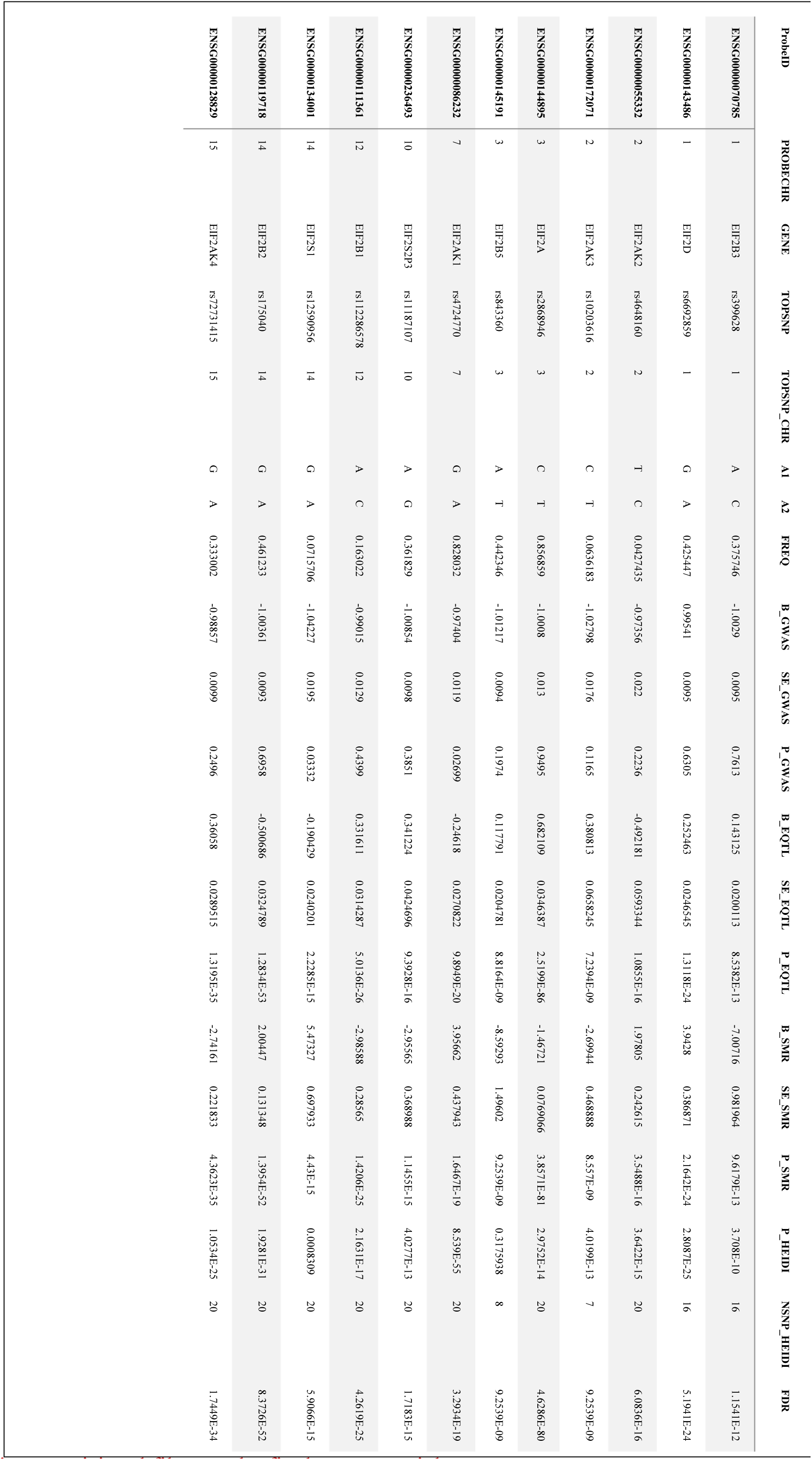
SMR/HEIDI results using the GWAS data on BD.

Since the ISR’s central regulatory switch lies in eIF2 ternary complex formation, we chose eIF2-related genes as exposure.

First, we conducted the SMR analysis to integrate the GWAS and blood eQTL data to identify the most relevant genes whose expression in blood was significantly associated with the trait of BD. Only one gene passed the SMR test. In chromosome 3, EIF2B5 gene passed both the SMR and HEIDI tests and thus were significantly associated with the trait of BD because of pleiotropy or causality.

Next, we conducted the COLOC analysis to integrate the GWAS and blood eQTL data of the genes that passed the SMR test and assess whether the genes were colocalized with the trait of BD. It can be seen from the data in Table 2 that there is strong support for colocalization between the trait and the EIF2B5 gene that passed both SMR and HEIDI tests.

**Table 2.**
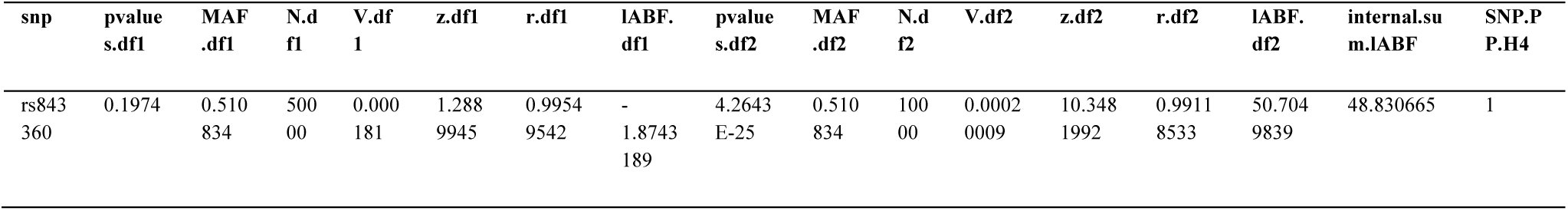
COLOC results of the genes that passed the SMR test.

### 3.2 Alt Drug Target

#### 3.2.1 Using PPI networks for alternative drug targets search

The assumption here is that the most likely alternative targets are either directly interacting with EIF2B5 or somewhere in the same PPI network. In this example, we consider only genes that were found to interact with EIF2B5 via direct protein-protein interactions, and require that those interacting proteins should also be druggable.

The thousands of genes are classified with regard to their druggability by (Finan et al., 2017), where the Tier 1 category refers to approved drugs or those in clinical testing while for other tier categories the druggability confidence drops in order Tier 2 and then Tier 3.

Table 3 shows the results obtained from data on the druggable alternative genes.

**Table 3.** Druggable alternative genes.

| g1.name | p1.uniprot_id | p2.uniprot_id | g2.name | g2.druggability_tier |
| --- | --- | --- | --- | --- |
| EIF2B5 | Q13144 | Q9P2K8 | EIF2AK4 | Tier 1 |
| EIF2B5 | Q13144 | P49841 | GSK3B | Tier 1 |
| EIF2B5 | Q13144 | Q9H1Y3 | OPN3 | Tier 3B |

#### 3.2.2 Distinguishing vertical and horizontal pleiotropy for SNP-protein associations

A key MR assumption is that the genetic variant (SNP) is only related to the outcome of interest through the exposure under study (the “exclusion restriction” assumption). This assumption is potentially violated under horizontal pleiotropy, where a SNP is associated with multiple phenotypes (e.g. proteins) independently of the exposure of interest. In contrast, vertical pleiotropy, where a SNP is associated with multiple phenotypes on the causal pathway to the outcome, does not violate the “exclusion restriction criterion” of MR.

Here, by integrating SNP-protein associations with biological pathway and protein-protein interaction (PPI) information retrieved from EpiGraphDB, we have developed an approach to assess potential horizontal pleiotropy. For a SNP associated with a group of proteins, we check the number of biological pathways and PPIs that are shared across this group of proteins. If these proteins are mapped to the same biological pathway and/or a PPI exists between them, then the SNP is more likely to act through vertical pleiotropy and therefore be a valid instrument for MR.

From the graph above we can see that P49841 and Q9P2K8 have same biological pathways, the table below illustrates the specific pathways that how they interact.

Then we complement the information given by pathway ontologies with data of interactions between proteins. For each pair of proteins, we retrieve shared protein-protein interactions, either a direct interaction or an interaction where there’s one mediator protein in the middle. As can be seen from the figure 3, for each pair of proteins, there are shared protein-protein interactions.

**Figure 2.**
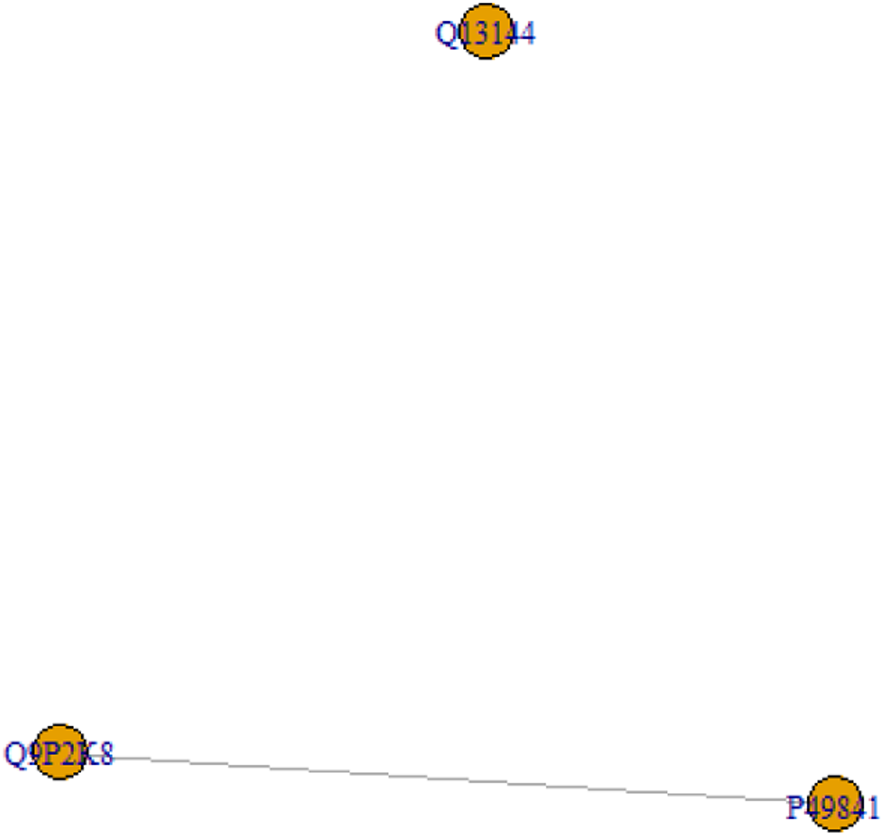
pathway_protein_graph.

**Figure 3.**
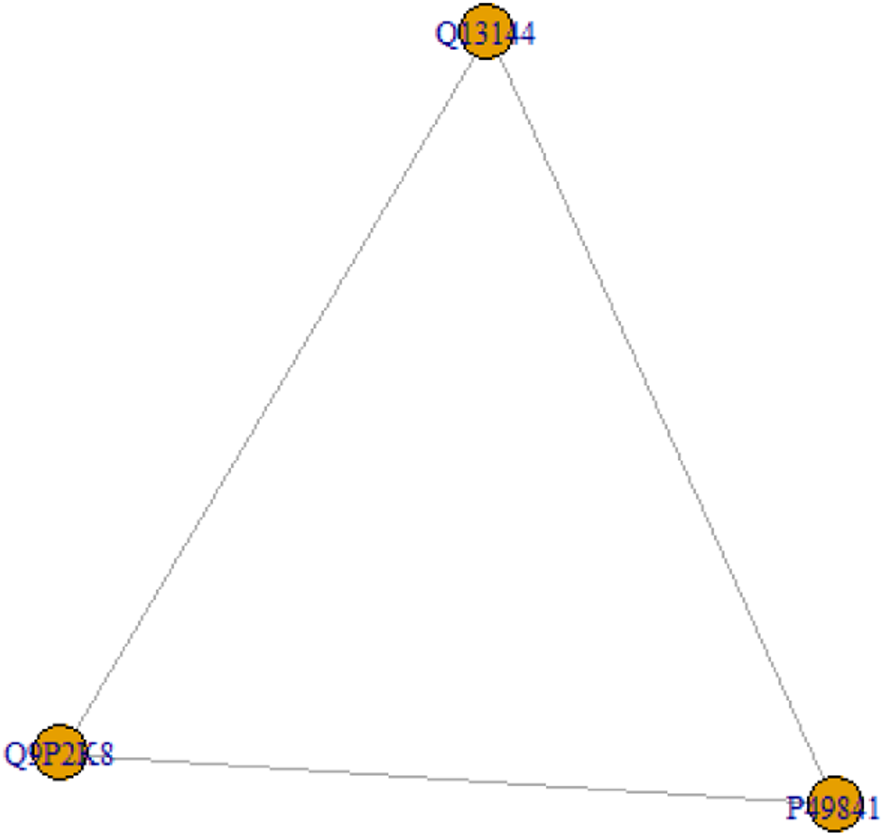
ppi_protein_graph.

Overall, these results indicate that our study will not violate under horizontal pleiotropy.

#### 3.2.3 Using Mendelian randomization results for causal effect estimation

The next step is to find out whether any of these genes have a comparable and statistically plausible effect on BD. For further analysis we select the gene of interest (EIF2B5) as well as its interacting genes with Tier 1 druggability.

**Figure 4.**
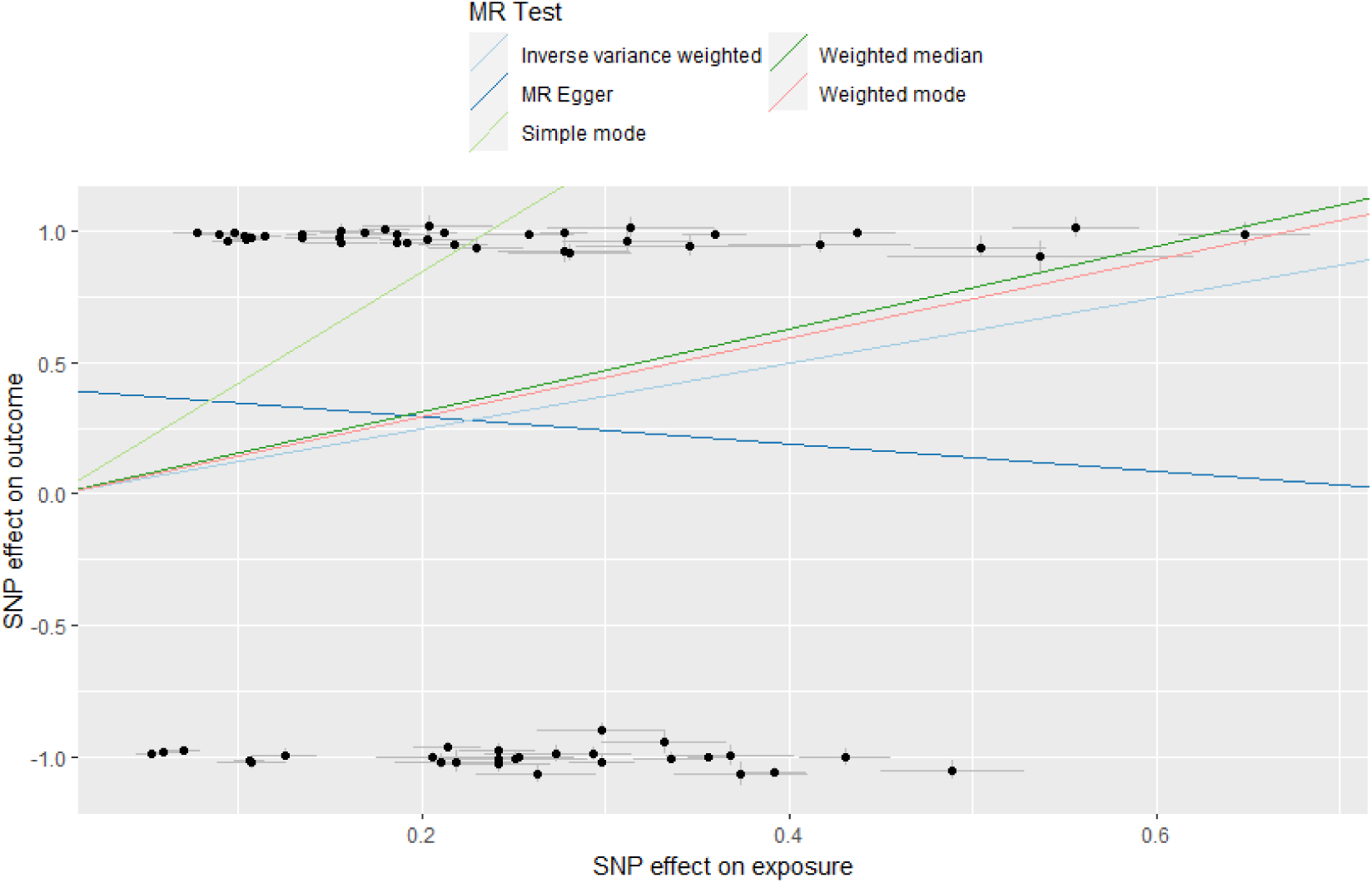
Scatter plots of the association of EIF2B5 and BD.

**Figure 5.**
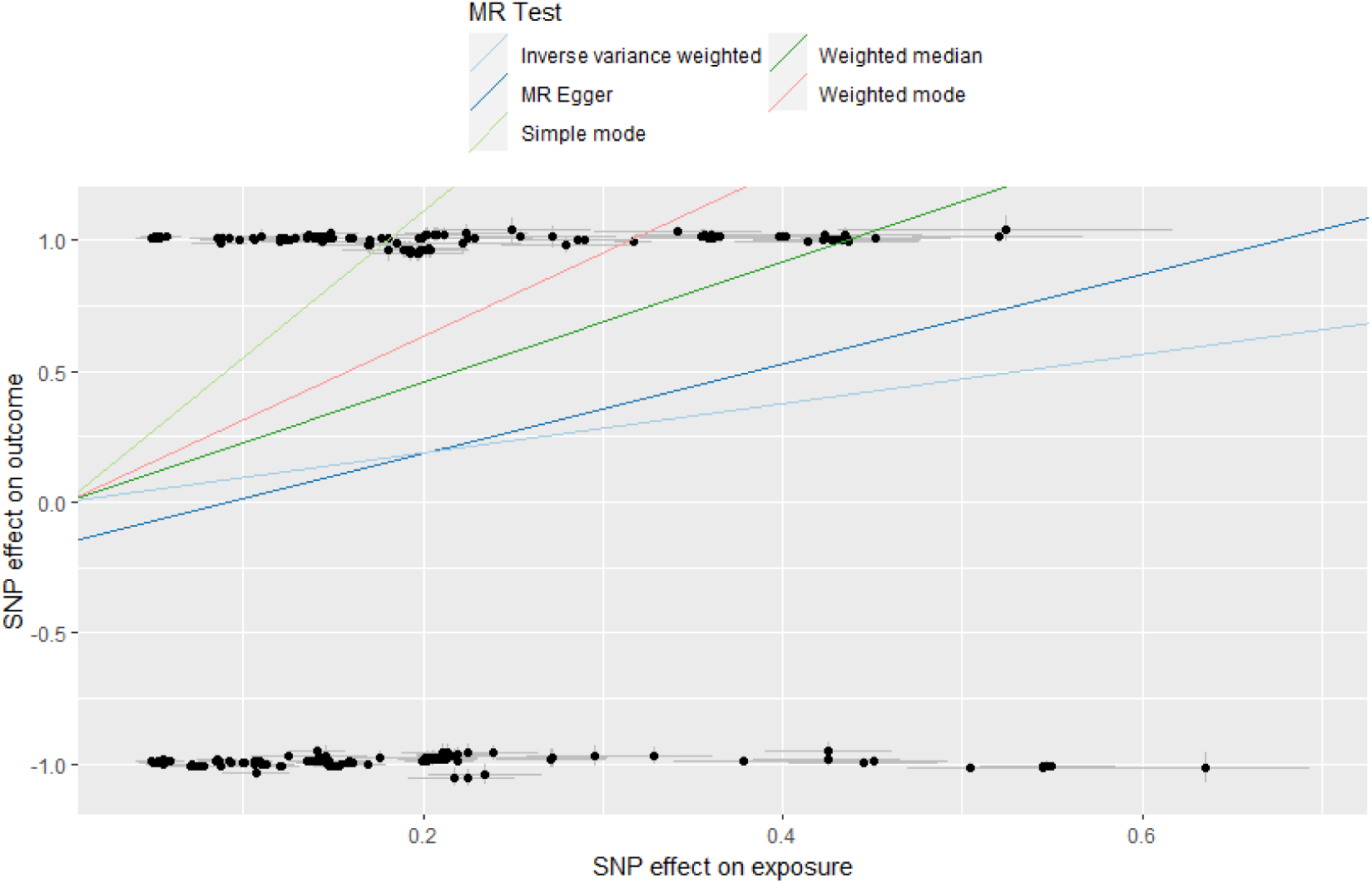
Scatter plots of the association of EIF2AK4 and BD.

**Figure 6.**
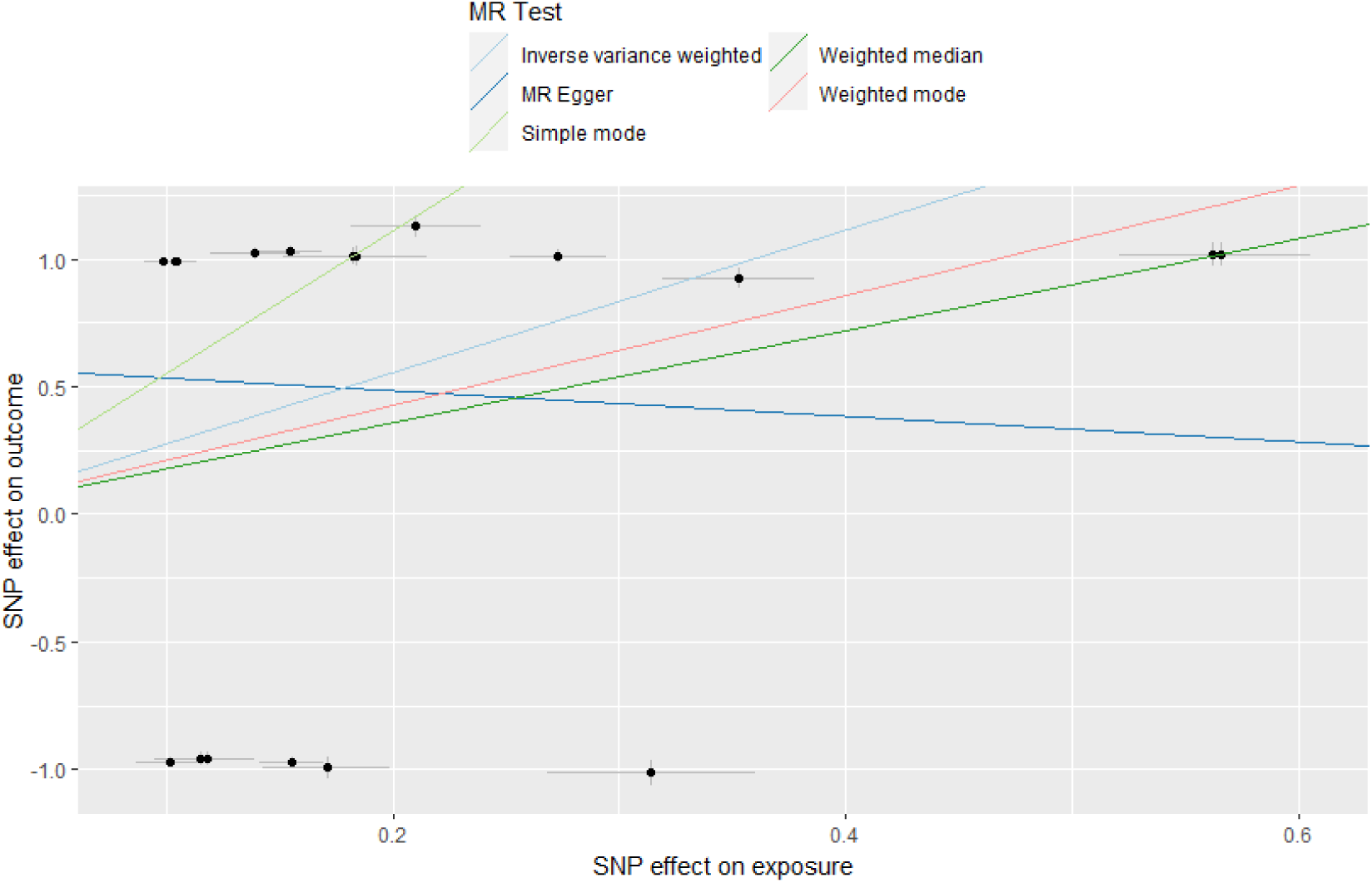
Scatter plots of the association of GSK3B and BD.

It is worth noting that there were also differences in the ratios of MR test. According to Burgess et al. (2023); Burgess and Thompson (2017), due to the heterogeneity test highlighting the existence of heterogeneity (P < 0.05), we give priority to using a random effect model of IVW and/or Weighted median method. The results, as shown in Table 5, indicate that a causal association was found between EIF2B5, EIF2AK4, GSK3B and BD.

**Table 4.** Specific pathways that how proteins interact.

| node.name | node.id | node.url |
| --- | --- | --- |
| Cellular responses to external stimuli | R-HSA-8953897 | https://reactome.org/PathwayBrowser/#/R-HSA-8953897 |
| Cellular responses to stress | R-HSA-2262752 | https://reactome.org/PathwayBrowser/#/R-HSA-2262752 |

**Table 5.**
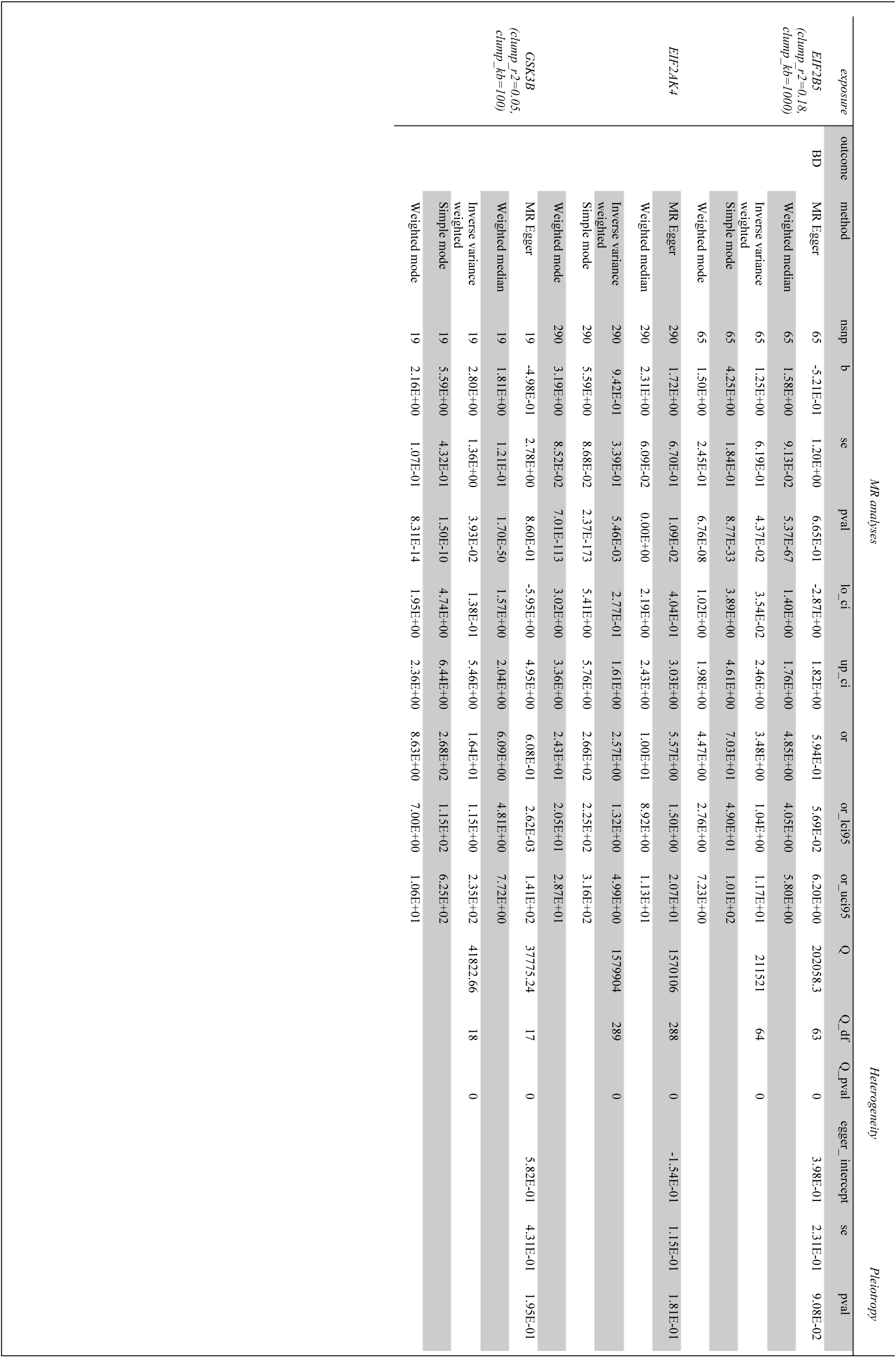
MR analysis of putative causal BD genes and bipolar disorder.

## 4 Discussion

The results of this study show that EIF2B5 is the potential disease-causing gene of BD, EIF2AK4 and GSK3B may become alternative drug targets for this disease.

Perhaps the most interesting finding is early research regarding EIF2B5 mainly aimed to study the roles of its expression in few cancerous tissue(Hou et al., 2020; Jiao et al., 2018; Xue et al., 2014; Yang et al., 2014), multiple sclerosis(Zahoor et al., 2017), and vanishing white matter (VWM) (Dietrich et al., 2005)syndrome. Importantly, the eIF2B complex plays a central role in the cellular integrated stress response (ISR). Stress-dependent kinase activation leads to phosphorylation of eIF2, which binds eIF2B more tightly and reduces overall protein synthesis. Previous research has established that white matter microstructure alterations in bipolar disorder(Adler et al., 2004; Bellani & Brambilla, 2011; Bellani et al., 2012; López-Larson et al., 2002; Yurgelun-Todd et al., 2007).

However, eukaryotic translation initiation factor 2B (EIF2B) gene family encoding the five subunits of eIF2B complex-α, β, γ, δ and ε respectively. This finding supports previous research into this brain area which links EIF2B5 and BD.

The ISR responds to a variety of different stress conditions that lead to alterations in cellular homeostasis. In metazoans, these stresses are sensed by at least four different kinases that each phosphorylate Ser51 in eIF2a to activate the ISR(Wek, 2018). One of them, GCN2(gene name EIF2AK4) contains a regulatory domain homologous with histidyl-tRNA synthetase and when amino acids are scarce, binding of deacylated His-tRNA triggers its activation (Harding et al., 2000; Hinnebusch, 2005). However, the precise mechanism(s) underlying these vastly different activation modalities remain unknown. But it is well known that the HPA axis is closely related to stress and AgRP neurons(Cleber Gama de Barcellos Filho et al., 2021; Perry et al., 2015; Perry et al., 2019). A possible explanation for this might further support the idea of Yuan et al. (2023): hypothalamic AgRP neurons are involved in the process of leucine deficiency improving depressive-like behavior induced by CRS, and the amino acid sensor general control non-derepressible 2 (GCN2) mediates this behavioral effect of AgRP neurons.

When it comes to GSK3B, Chen et al. (2014) showed that published data on the association between GSK3B –50C/T (rs334558) and bipolar disorder are inconclusive. This differs from the findings presented here. It could be argued that the positive results were due to the relatively much bigger sample size and a multiple SNPs-based approach can provide a more objective evaluation. Despite its exploratory nature, this study offers some insight into GSK3B, which is not only implicated in the pathophysiology of Alzheimer disease (Mendes et al., 2009), Parkinson disease (Wider et al., 2011), schizophrenia (Chen et al., 2020), and depressive disorders (Levchenko et al., 2018).

The generalisability of these results is subject to certain limitations. Firstly, the SMR method cannot distinguish pleiotropic genes from the causative genes. The association between prioritized genes and BD needs to be validated by follow-up functional studies. Moreover, BD cohorts in our data were collected in Europe, North America and Australia, so these findings cannot be extrapolated to all patients.

## 5 Conclusion

This study set out to examine the relationship between ISR and BD, also, assess the feasibility of some genes’ druggability. The most obvious finding to emerge from this study is that we established causal relationships between genetic variants, candidate genes, and BD risk. The insights gained from this study may be of assistance to cure this disease.

## 7 Author Contributions

ZT designed this study, analyzed data, and wrote the draft of the manuscript. Prof.Deng huihua discussed and reviewed the manuscript critically. All authors contributed to the article and approved the submitted version.

## 8 Acknowledgments

The authors thank Yang Lab for the user-friendly SMR software. We also appreciate consortia such as the PGC, GTEx and eQTLGen for providing publicly available data.

## 9 Data Availability Statement

The summary-level data of the GWAS on BD was publicly available from the Psychiatric Genomics Consortium (https://figshare.com/articles/dataset/bip2021_noUKBB/22564402/1). The eQTL dataset was publicly available from the eQTLGen consortium (https://www.eqtlgen.org/). V8 release of the GTEx eQTL/sQTL summary data can be downloaded from https://yanglab.westlake.edu.cn/software/smr/#DataResource.

## Reference

1. Adler, C. M., Holland, S. K., Schmithorst, V., Wilke, M., Weiss, K. L., Pan, H., & Strakowski, S. M. (2004). Abnormal frontal white matter tracts in bipolar disorder: a diffusion tensor imaging study. Bipolar Disorders, 6(3), 197–203. 10.1111/j.1399-5618.2004.00108.x

2. Atkin, J. D., Farg, M. A., Walker, A. K., McLean, C., Tomas, D., & Horne, M. K. (2008). Endoplasmic reticulum stress and induction of the unfolded protein response in human sporadic amyotrophic lateral sclerosis. Neurobiology of Disease, 30(3), 400–407. 10.1016/j.nbd.2008.02.009

3. Bellani, M., & Brambilla, P. (2011). Diffusion imaging studies of white matter integrity in bipolar disorder. Epidemiology and Psychiatric Sciences, 20(2), 137–140. 10.1017/s2045796011000229

4. Bellani, M., Perlini, C., Ferro, A., Cerruti, S., Rambaldelli, G., Isola, M., Cerini, R., Dusi, N., Andreone, N., Balestrieri, M., Mucelli, R. P., Tansella, M., & Brambilla, P. (2012). White matter microstructure alterations in bipolar disorder. Functional Neurology, 27(1), 29–34. https://pubmed.ncbi.nlm.nih.gov/22687164/

5. Buffington, S. A., Huang, W., & Costa-Mattioli, M. (2014). Translational Control in Synaptic Plasticity and Cognitive Dysfunction. In S. E. Hyman (Ed.), Annual Review of Neuroscience, Vol 37 (Vol. 37, pp. 17–38). Annual Reviews. 10.1146/annurev-neuro-071013-014100

6. Burgess, S., Davey Smith, G., Davies, N., Dudbridge, F., Gill, D., Glymour, M., Hartwig, F., Kutalik, Z., Holmes, M., Minelli, C., Morrison, J., Pan, W., Relton, C., & Theodoratou, E. (2023). Guidelines for performing Mendelian randomization investigations: update for summer 2023 [version 3; peer review: 2 approved]. Wellcome Open Research, 4(186). 10.12688/wellcomeopenres.15555.3

7. Burgess, S., & Thompson, S. G. (2017). Interpreting findings from Mendelian randomization using the MR-Egger method. European Journal of Epidemiology, 32(5), 377–389. 10.1007/s10654-017-0255-x

8. Chang, R. C. C., Wong, A. K. Y., Ng, H.-K., & Hugon, J. (2002). Phosphorylation of eukaryotic initiation factor-2α (eIF2α) is associated with neuronal degeneration in Alzheimer’s disease. NeuroReport, 13(18), 2429–2432. 10.1097/00001756-200212200-00011

9. Chen, G. D., Tang, J., Yu, G. W., Chen, Y. P., Wang, L. C., & Zhang, Y. (2014). Meta-analysis demonstrates lack of association of the GSK3B-50C/T polymorphism with risk of bipolar disorder. Molecular Biology Reports, 41(9), 5711–5718. 10.1007/s11033-014-3441-x

10. Chen, Y., Hua, S., Wang, W. P., Fan, W. X., Tang, W., Zhang, Y., & Zhang, C. (2020). A comprehensive analysis of GSK3B variation for schizophrenia in Han Chinese individuals. Asian Journal of Psychiatry, 47, Article 101832. 10.1016/j.ajp.2019.10.012

11. Chou, A., Krukowski, K., Jopson, T., Zhu, P. J., Costa-Mattioli, M., Walter, P., & Rosi, S. (2017). Inhibition of the integrated stress response reverses cognitive deficits after traumatic brain injury. Proceedings of the National Academy of Sciences of the United States of America, 114(31), E6420–E6426. 10.1073/pnas.1707661114

12. Cleber Gama de Barcellos Filho, P., Campos Zanelatto, L., Amélia Aparecida Santana, B., Calado, R. T., & Rodrigues Franci, C. (2021). Effects chronic administration of corticosterone and estrogen on HPA axis activity and telomere length in brain areas of female rats. Brain Research, 1750, 147152. 10.1016/j.brainres.2020.147152

13. Consortium, T. G., Aguet, F., Anand, S., Ardlie, K. G., Gabriel, S., Getz, G. A., Graubert, A., Hadley, K., Handsaker, R. E., Huang, K. H., Kashin, S., Li, X., MacArthur, D. G., Meier, S. R., Nedzel, J. L., Nguyen, D. T., Segrè, A. V., Todres, E., Balliu, B., … Volpi, S. (2020). The GTEx Consortium atlas of genetic regulatory effects across human tissues. Science, 369(6509), 1318–1330. 10.1126/science.aaz1776

14. Costa-Mattioli, M., & Walter, P. (2020). The integrated stress response: From mechanism to disease. Science, 368(6489), 384-+, Article eaat5314. 10.1126/science.aat5314

15. de Kloet, E. R., Joëls, M., & Holsboer, F. (2005). Stress and the brain:: From adaptation to disease. Nature Reviews Neuroscience, 6(6), 463–475. 10.1038/nrn1683

16. de Kloet, E. R., Sibug, R. M., Helmerhorst, F. M., & Schmidt, M. (2005). Stress, genes and the mechanism of programming the brain for later life. Neuroscience and Biobehavioral Reviews, 29(2), 271–281. 10.1016/j.neubiorev.2004.10.008

17. *Diagnostic and statistical manual of mental disorders: DSM-5™, 5th ed*. (2013). American Psychiatric Publishing, Inc. 10.1176/appi.books.9780890425596

18. Dietrich, J., Lacagnina, M., Gass, D., Richfield, E., Mayer-Pröschel, M., Noble, M., Torres, C., & Pröschel, C. (2005). EIF2B5 mutations compromise GFAP + astrocyte generation in vanishing white matter leukodystrophy. Nature Medicine, 11(3), 277–283. 10.1038/nm1195

19. Emdin, C. A., Khera, A. V., & Kathiresan, S. (2017). Mendelian Randomization. JAMA, 318(19), 1925–1926. 10.1001/jama.2017.17219

20. Finan, C., Gaulton, A., Kruger, F. A., Lumbers, R. T., Shah, T., Engmann, J., Galver, L., Kelley, R., Karlsson, A., Santos, R., Overington, J. P., Hingorani, A. D., & Casas, J. P. (2017). The druggable genome and support for target identification and validation in drug development. Science Translational Medicine, 9(383), eaag1166. 10.1126/scitranslmed.aag1166

21. Giambartolomei, C., Vukcevic, D., Schadt, E. E., Franke, L., Hingorani, A. D., Wallace, C., & Plagnol, V. (2014). Bayesian Test for Colocalisation between Pairs of Genetic Association Studies Using Summary Statistics. Plos Genetics, 10(5), Article e1004383. 10.1371/journal.pgen.1004383

22. Gold, P. W., Goodwin, F. K., & Chrousos, G. P. (1988). CLINICAL AND BIOCHEMICAL MANIFESTATIONS OF DEPRESSION .1. RELATION TO THE NEUROBIOLOGY OF STRESS. New England Journal of Medicine, 319(6), 348–353. 10.1056/nejm198808113190606

23. Grande, I., Berk, M., Birmaher, B., & Vieta, E. (2016). Bipolar disorder [Review]. Lancet, 387(10027), 1561–1572. 10.1016/s0140-6736(15)00241-x

24. Gu, F., Chauhan, V., & Chauhan, A. (2015). Glutathione redox imbalance in brain disorders. Current Opinion in Clinical Nutrition and Metabolic Care, 18(1), 89–95. 10.1097/mco.0000000000000134

25. Harding, H. P., Novoa, I., Zhang, Y., Zeng, H., Wek, R., Schapira, M., & Ron, D. (2000). Regulated Translation Initiation Controls Stress-Induced Gene Expression in Mammalian Cells. Molecular Cell, 6(5), 1099–1108. 10.1016/S1097-2765(00)00108-8

26. Hemani, G., Zheng, J., Elsworth, B., Wade, K. H., Haberland, V., Baird, D., Laurin, C., Burgess, S., Bowden, J., Langdon, R., Tan, V. Y., Yarmolinsky, J., Shihab, H. A., Timpson, N. J., Evans, D. M., Relton, C., Martin, R. M., Davey Smith, G., Gaunt, T. R., & Haycock, P. C. (2018). The MR-Base platform supports systematic causal inference across the human phenome. eLife, 7, e34408. 10.7554/eLife.34408

27. Hinnebusch, A. G. (2005). TRANSLATIONAL REGULATION OF GCN4 AND THE GENERAL AMINO ACID CONTROL OF YEAST. Annual Review of Microbiology, 59(1), 407–450. 10.1146/annurev.micro.59.031805.133833

28. Hoozemans, J. J. M., van Haastert, E. S., Eikelenboom, P., de Vos, R. A. I., Rozemuller, J. M., & Scheper, W. (2007). Activation of the unfolded protein response in Parkinson’s disease. Biochemical and Biophysical Research Communications, 354(3), 707–711. 10.1016/j.bbrc.2007.01.043

29. Hou, L., Jiao, Y., Li, Y. Q., Luo, Z. P., Zhang, X. Y., Pan, G. Q., Zhao, Y. C., Yang, Z. Y., & He, M. (2020). Low EIF2B5 expression predicts poor prognosis in ovarian cancer. Medicine, 99(5), 6, Article e18666. 10.1097/md.0000000000018666

30. Hwang, K. D., Bak, M. S., Kim, S. J., Rhee, S., & Lee, Y. S. (2017). Restoring synaptic plasticity and memory in mouse models of Alzheimer’s disease by PKR inhibition. Molecular Brain, 10, 10, Article 57. 10.1186/s13041-017-0338-3

31. Ishimura, R., Nagy, G., Dotu, I., Chuang, J. H., & Ackerman, S. L. (2016). Activation of GCN2 kinase by ribosome stalling links translation elongation with translation initiation. eLife, 5, 22, Article e14295. 10.7554/eLife.14295

32. Jiao, Y., Fu, Z., Li, Y. Q., Meng, L. Y., & Liu, Y. H. (2018). High EIF2B5 mRNA expression and its prognostic significance in liver cancer: a study based on the TCGA and GEO database. Cancer Management and Research, 10, 6003–6014. 10.2147/cmar.S185459

33. Kanner, A. M. (2012). Can neurobiological pathogenic mechanisms of depression facilitate the development of seizure disorders? Lancet Neurology, 11(12), 1093–1102. 10.1016/s1474-4422(12)70201-6

34. Kulak, A., Steullet, P., Cabungcal, J. H., Werge, T., Ingason, A., Cuenod, M., & Do, K. Q. (2013). Redox Dysregulation in the Pathophysiology of Schizophrenia and Bipolar Disorder: Insights from Animal Models. Antioxidants & Redox Signaling, 18(12), 1428–1443. 10.1089/ars.2012.4858

35. Lackovic, M., Rovcanin, B., Pantovic, M., Ivkovic, M., Petronijevic, N., & Damjanovic, A. (2013). ASSOCIATION OF OXIDATIVE STRESS WITH THE PATHOPHYSIOLOGY OF DEPRESSION AND BIPOLAR DISORDER. Archives of Biological Sciences, 65(1), 369–373. 10.2298/abs1301369l

36. Leboyer, M., Soreca, I., Scott, J., Frye, M., Henry, C., Tamouza, R., & Kupfer, D. J. (2012). Can bipolar disorder be viewed as a multi-system inflammatory disease? Journal of Affective Disorders, 141(1), 1–10. 10.1016/j.jad.2011.12.049

37. Levchenko, A., Losenkov, I. S., Vyalova, N. M., Simutkin, G. G., Bokhan, N. A., Wilffert, B., Loonen, A. J. M., & Ivanova, S. A. (2018). The functional variant rs334558 of GSK3B is associated with remission in patients with depressive disorders. Pharmacogenomics & Personalized Medicine, 11, 121–126. 10.2147/pgpm.S171423

38. Liu, Y., Elsworth, B., Erola, P., Haberland, V., Hemani, G., Lyon, M., Zheng, J., Lloyd, O., Vabistsevits, M., & Gaunt, T. R. (2020). EpiGraphDB: a database and data mining platform for health data science. Bioinformatics, 37(9), 1304–1311. 10.1093/bioinformatics/btaa961

39. López-Larson, M. P., DelBello, M. P., Zimmerman, M. E., Schwiers, M. L., & Strakowski, S. M. (2002). Regional prefrontal gray and white matter abnormalities in bipolar disorder. Biological Psychiatry, 52(2), 93–100, Article Pii s0006-3223(02)01350-1. 10.1016/s0006-3223(02)01350-1

40. Mayer, F. L., Sánchez-León, E., & Kronstad, J. W. (2018). A chemical genetic screen reveals a role for proteostasis in capsule and biofilm formation by Cryptococcus neoformans. Microbial Cell, 5(11), 495–510. 10.15698/mic2018.11.656

41. Mendes, C. T., Mury, F. B., Moreira, E. D., Alberto, F. L., Forlenza, O. V., Dias-Neto, E., & Gattaz, W. F. (2009). Lithium reduces Gsk3b mRNA levels: implications for Alzheimer Disease. European Archives of Psychiatry and Clinical Neuroscience, 259(1), 16–22. 10.1007/s00406-008-0828-5

42. Moon, S. L., Sonenberg, N., & Parker, R. (2018). Neuronal Regulation of eIF2α Function in Health and Neurological Disorders. Trends in Molecular Medicine, 24(6), 575–589. 10.1016/j.molmed.2018.04.001

43. Perry, R. J., Lee, S., Ma, L., Zhang, D., Schlessinger, J., & Shulman, G. I. (2015). FGF1 and FGF19 reverse diabetes by suppression of the hypothalamic–pituitary–adrenal axis. Nature communications, 6(1), 6980. 10.1038/ncomms7980

44. Perry, R. J., Resch, J. M., Douglass, A. M., Madara, J. C., Rabin-Court, A., Kucukdereli, H., Wu, C., Song, J. D., Lowell, B. B., & Shulman, G. I. (2019). Leptin’s hunger-suppressing effects are mediated by the hypothalamic–pituitary–adrenocortical axis in rodents. Proceedings of the National Academy of Sciences, 116(27), 13670–13679. doi:10.1073/pnas.1901795116

45. Prusty, B. K., Gulve, N., Govind, S., Krueger, G. R. F., Feichtinger, J., Larcombe, L., Aspinall, R., Ablashi, D. V., & Toro, C. T. (2018). Active HHV-6 Infection of Cerebellar Purkinje Cells in Mood Disorders. Frontiers in Microbiology, 9, Article 1955. 10.3389/fmicb.2018.01955

46. Rosenblat, J. D., & McIntyre, R. S. (2015). Are medical comorbid conditions of bipolar disorder due to immune dysfunction? Acta Psychiatrica Scandinavica, 132(3), 180–191. 10.1111/acps.12414

47. Rowland, T. A., & Marwaha, S. (2018). Epidemiology and risk factors for bipolar disorder. Therapeutic Advances in Psychopharmacology, 8(9), 251–269. 10.1177/2045125318769235

48. Sen, T., Gupta, R., Kaiser, H., & Sen, N. (2017). Activation of PERK Elicits Memory Impairment through Inactivation of CREB and Downregulation of PSD95 After Traumatic Brain Injury. Journal of Neuroscience, 37(24), 5900–5911. 10.1523/jneurosci.2343-16.2017

49. Smith, H. L., & Mallucci, G. R. (2016). The unfolded protein response: mechanisms and therapy of neurodegeneration. Brain, 139, 2113–2121. 10.1093/brain/aww101

50. Sullivan, P. (2023). bip2021_noUKBB. figshare. 10.6084/m9.figshare.22564402.v1

51. Tible, M., Liger, F. M., Schmitt, J., Giralt, A., Farid, K., Thomasseau, S., Gourmaud, S., Paquet, C., Reig, L. R., Meurs, E., Girault, J. A., & Hugon, J. (2019). PKR knockout in the 5xFAD model of Alzheimer’s disease reveals beneficial effects on spatial memory and brain lesions. Aging Cell, 18(3), 11, Article e12887. 10.1111/acel.12887

52. Vieta, E., Salagre, E., Grande, I., Carvalho, A. F., Fernandes, B. S., Berk, M., Birmaher, B., Tohen, M., & Suppes, T. (2018). Early Intervention in Bipolar Disorder. American Journal of Psychiatry, 175(5), 411–426. 10.1176/appi.ajp.2017.17090972

53. Võsa, U., Claringbould, A., Westra, H.-J., Bonder, M. J., Deelen, P., Zeng, B., Kirsten, H., Saha, A., Kreuzhuber, R., Yazar, S., Brugge, H., Oelen, R., de Vries, D. H., van der Wijst, M. G. P., Kasela, S., Pervjakova, N., Alves, I., Favé, M.-J., Agbessi, M., … i, Q. T. L. C. (2021). Large-scale cis– and trans-eQTL analyses identify thousands of genetic loci and polygenic scores that regulate blood gene expression. Nature Genetics, 53(9), 1300–1310. 10.1038/s41588-021-00913-z

54. Wang, G., Sarkar, A., Carbonetto, P., & Stephens, M. (2020). A Simple New Approach to Variable Selection in Regression, with Application to Genetic Fine Mapping. Journal of the Royal Statistical Society Series B: Statistical Methodology, 82(5), 1273–1300. 10.1111/rssb.12388

55. Wek, R. C. (2018). Role of eIF2α kinases in translational control and adaptation to cellular stress. Cold Spring Harbor perspectives in biology, 10(7), a032870. 10.1101/cshperspect.a032870

56. Wider, C., Vilariño-Güell, C., Heckman, M. G., Jasinska-Myga, B., Ortolaza-Soto, A. I., Diehl, N. N., Crook, J. E., Cobb, S. A., Bacon, J. A., Aasly, J. O., Gibson, J. M., Lynch, T., Uitti, R. J., Wszolek, Z. K., Farrer, M. J., & Ross, O. A. (2011). SNCA, MAPT, and GSK3B in Parkinson disease: a gene-gene interaction study. European Journal of Neurology, 18(6), 876–881. 10.1111/j.1468-1331.2010.03297.x

57. Wu, Y., Broadaway, K. A., Raulerson, C. K., Scott, L. J., Pan, C., Ko, A., He, A., Tilford, C., Fuchsberger, C., Locke, A. E., Stringham, H. M., Jackson, A. U., Narisu, N., Kuusisto, J., Pajukanta, P., Collins, F. S., Boehnke, M., Laakso, M., Lusis, A. J., … Mohlke, K. L. (2019). Colocalization of GWAS and eQTL signals at loci with multiple signals identifies additional candidate genes for body fat distribution. Human Molecular Genetics, 28(24), 4161–4172. 10.1093/hmg/ddz263

58. Xue, D., Lu, M., Gao, B., Qiao, X., & Zhang, Y. (2014). Screening for transcription factors and their regulatory small molecules involved in regulating the functions of CL1-5 cancer cells under the effects of macrophage-conditioned medium. Oncol Rep, 31(3), 1323–1333. 10.3892/or.2013.2937

59. Yang, S., Zhang, H., Guo, L., Zhao, Y., & Chen, F. (2014). Reconstructing the coding and non–coding RNA regulatory networks of miRNAs and mRNAs in breast cancer. Gene, 548(1), 6–13. 10.1016/j.gene.2014.06.010

60. Yuan, F., Wu, S., Zhou, Z., Jiao, F., Yin, H., Niu, Y., Jiang, H., Chen, S., & Guo, F. (2023). Leucine deprivation results in antidepressant effects via GCN2 in AgRP neurons. Life Metabolism, 2(1). 10.1093/lifemeta/load004

61. Yurgelun-Todd, D. A., Silveri, M. M., Gruber, S. A., Rohan, M. L., & Pimentel, P. J. (2007). White matter abnormalities observed in bipolar disorder: a diffusion tensor imaging study. Bipolar Disorders, 9(5), 504–512. 10.1111/j.1399-5618.2007.00395.x

62. Zahoor, I., Haq, E., & Asimi, R. (2017). Multiple Sclerosis and EIF2B5: A Paradox or a Missing Link. In A. A. A. Asea, F. Geraci, & P. Kaur (Eds.), Multiple Sclerosis: Bench to Bedside: Global Perspectives on a Silent Killer (Vol. 958, pp. 57-64). Springer International Publishing Ag. 10.1007/978-3-319-47861-6_5

63. Zhu, Z., Zhang, F., Hu, H., Bakshi, A., Robinson, M. R., Powell, J. E., Montgomery, G. W., Goddard, M. E., Wray, N. R., Visscher, P. M., & Yang, J. (2016). Integration of summary data from GWAS and eQTL studies predicts complex trait gene targets. Nature Genetics, 48(5), 481–487. 10.1038/ng.3538

64. Zhuang, J., Li, F., Liu, X., Liu, Z. P., Lin, J. X., Ge, Y. H., Kaminski, J. M., Summers, J. B., Wang, Z. C., Ge, J., & Yu, K. M. (2009). Lithium chloride protects retinal neurocytes from nutrient deprivation by promoting DNA non-homologous end-joining. Biochemical and Biophysical Research Communications, 380(3), 650–654. 10.1016/j.bbrc.2009.01.162

